# On the progression of COVID19 in Portugal: A comparative analysis of active cases using non-linear regression

**DOI:** 10.1101/2020.05.02.20088856

**Authors:** Ana Milhinhos, Pedro M. Costa

## Abstract

Portugal has been portrayed as a relatively successful case in the control of the COVID-19’s March 2020 outbreak in Europe due to the timely confinement measures taken. As other European Union member states, Portugal is now preparing the phased loosening of the confinement measures, starting in the beginning of May. Even so, the current data, albeit showing at least a reduction in infection rates, renders difficult to forecast scenarios in the imminent future. Using South Korea data as scaffold, which is becoming a paradigmatic case of recovery following a high number of infected people, we fitted Portuguese data to biphasic models using non-linear regression and compared the two countries. The results, which suggest good fit, show that recovery in Portugal can be much slower than anticipated, with a very high percentage of active cases (over 50%) remaining still active even months after the projected end of mitigation measures. This, together with the unknown number of asymptomatic carriers, may increase the risk of a much slower recovery if not of new outbreaks. Europe and elsewhere must consider this contingency when planning the relief of containment measures.

---

The first documented case of COVID-19 infection in Portugal dates March 2, 2020. Motivated by the rapid progression in other countries, especially in neighbouring Spain (see Kinross et al., 2020; Spiteri et al., 2020), the country moved swiftly to control dissemination by shutting down many public services and imposing strict confinement measures (see for instance Mahase, 2020). These date from mid-March and, by the time the present work is prepared, the Portuguese government and competent health authorities are planning the phased cessation of these measures, in alignment with the European Union. It has been consensual that the spreading of COVID-19 in Portugal, with respect to number of infected people, fatalities and ICU internments, is reaching a plateau, making it a potential case of success, as by mid-April fatalities were kept below 1,000 and the healthcare system did not attain saturation. Now is a crucial time to know what to expect from the progression of the disease in the country and how safe it is to begin the relief of confinement measures. However, the available data is still insufficient to draw solid forecasts even on short-term. Indeed, at this stage, epidemiological SIR (‘susceptible’, ‘infected’ and ‘recovered’) models are difficult to produce in Portugal and elsewhere.

Lessons can be learned from the few countries already clearing the pandemic. The Republic of Korea is a key case study not just due to the overall positive progress but also because the country implemented strict confinement measures, imposed timely limitations to in-bound travelling and closed public services like schools. Also, South Korea endured a high number of total infections (which offers statistical significance), albeit a relatively low mortality rate, estimated at 0.9% by mid-March, when cases totalled almost 8,000, according to the Republic of Korea COVID-19 National Emergency Response Center (2020). Still, the basic reproduction number (R_0_) has been estimated at 1.5 ± 0.1 (Shim et al., 2020), therefore within the magnitude of the influenza outbreak in 1918 (Ferguson et al., 2006). Consequently, there are significant similarities between countries albeit likely differences in public behaviour, awareness or susceptibility. We therefore aimed at modelling the progression of active cases in Portugal (up to April 19) by means of non-linear regression using Korean data as scaffold.

We used a four-parameter log-logistic model to estimate the maximum number of infections in Portugal, which, according to the current data, will surpass 25,500. The same model yielded a maximum of about 1,000 daily fatalities by day 116 (June 24) since the first cases were reported (March 2), a date that landmarks the beginning of the outbreak in Portugal (Fig. 1A). The current mortality rate is 3.5 *%* (as per April 19), well below Italy, with over 10% (Rubino et al., 2020). One of the most positive signs for COVID-19 control in Portugal has been considered the reduced percentage of daily cases (Fig. 1B). This information must, nonetheless, be interpreted with caution as reported new cases are highly variable, in part due to increased testing. Active cases were then fitted to a five-parameter log-Gaussian distribution, as described by Martin-Betancor et al. (2015), a biphasic asymmetric model. In fact, the recovery rates in Portugal are low (only 610 cases by April 19), seemingly accordant to reports from elsewhere. At this stage, Portugal as yet to reach the peak of active cases, which means that data cannon be fitted to the descending phase of the curve. Still, the fit was nearly perfect to the ascending phase (Fig. 1C). Moreover, Korean data also fitted the same model perfectly, yielding, as expected, slower recovery than infection rates (Fig. 1D). By juxtaposing the two models and expanding them to a full-year timeframe, the differences between the two countries become evident (Fig. 1E). At day 50, South Korea reported 3591 active cases whereas the model estimated 3653 cases (half of the 7307 projected maximum), with the real maximum being 7293, which, again, shows the good fit of the model. Portugal may only reach 50% of recoveries after 140 days. The different shapes of curves, reflected in differential parametrization of models (Table 1) should reflect not only the number of cases but also different rates of recovery.

**Table 1.** Summary of parameter estimates for the fitting of active COVID-19 cases in Portugal and the Republic of Korea by April 19, 2020. The fitted model consisted of a five-parameter log-Gaussian distribution, a biphasic response model. Parameters were obtained through least squares estimation, using the package ‘drc’. All statistics were performed using R 3.5 (Ihaka and Gentleman, 1996). The Portuguese data (March 2 – April 19) were compiled from the official daily reports on COVID-19 provided by the General Directorate for Health, available at https://covid19.min-saude.pt/ponto-de-situacao-atual-em-portugal/. Data from South Korea (February 15 – April 19) were retrieved from Worldometer (https://www.worldometers.info/coronavirus/). Data is provided in Supplementary information.

|  | Parameter | Estimate | Standard error | *t*-value | *p*-value |
| --- | --- | --- | --- | --- | --- |
| Portugal |  |  |  |  |  |
|  | *b* | 0.60 | 1.53E-01 | 3.9315 | 0.0002885 |
|  | *c* | 51.52 | 4.76E+01 | 1.0822 | 0.2849116 |
|  | *d*<sup>1</sup> | 21,559.00 | 1.45E+03 | 14.9038 | < 2.2E-16 |
|  | *e* | 69.50 | 1.30E+01 | 5.3548 | 2.799E-06 |
|  | *f* | 2.40 | 3.98E-01 | 6.0222 | 2.894E-07 |
| Republic of Korea |  |  |  |  |  |
|  | *b* | 0.48 | 1.49E-02 | 32.185 | <2E-16 |
|  | *c* | -66.75 | 1.33E+02 | -0.502 | 0.6176 |
|  | *d*<sup>1</sup> | 7,306.07 | 1.25E+02 | 58.253 | <2E-16 |
|  | *e* | 27.69 | 1.92E-01 | 144.09 | <2E-16 |
|  | *f* | 1.71 | 9.64E-02 | 17.706 | <2E-16 |

<sup>1</sup> parameter *d* is the predicted maximum

**Fig. 1.**
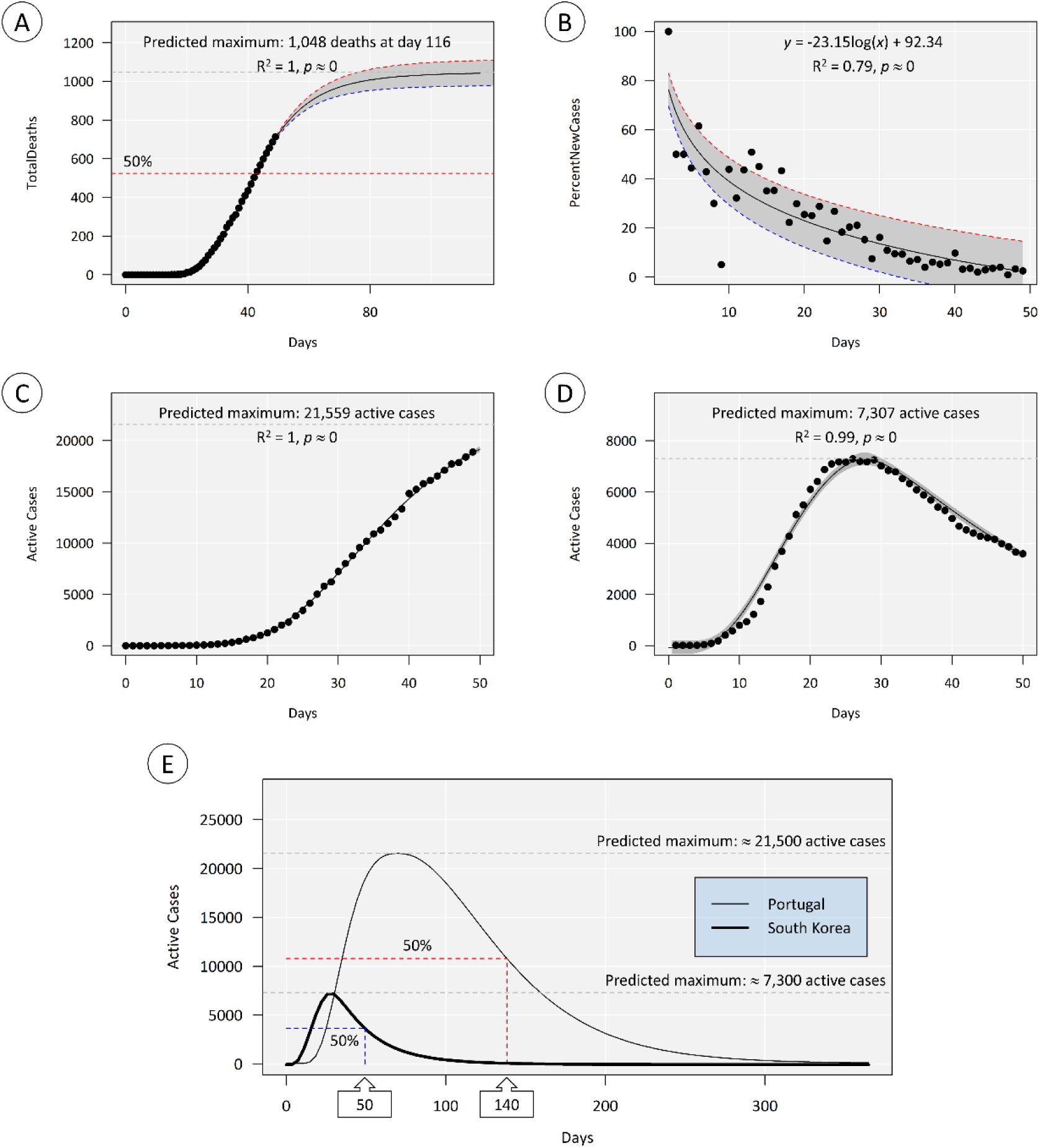
An overview of the evolution of COVID-19 in Portugal from March 1 (day 0) to April 19 (day 49) 2020. A) Cumulative number of deaths fitted to a log-logistic model. Scale was extended to highlight the quality of fit and predicted asymptotic limit (≈ 1,000 deaths). B) A simple log-linear regression for the percentage of daily new cases (infected subjects) relatively to cumulative new cases. C) Total active cases (i.e. total cases excluding deaths and recoveries) fitted to a log-Gaussian (asymmetric) model with an estimated maximum at ≈ 21,500 cases, highlighting the near perfect fit to the growth phase of the model. D) Active cases reported in the Republic of Korea between February 15 and April 19 fitted to log-Gaussian Model, as previous. The South Korean scenario already has sufficient data to fit both growth and decrease phases, again yielding a near perfect fit. E) Juxtaposition of predicted models (scaled to a full year from the first day of reported cases) for Portuguese and Korean data (log-Gaussian non-linear regression). The models highlighting maxima and half-maximal estimates (50% of cases recovered). Whereas south Korea already surpassed the estimate (as day 50 corresponds to April 4), in Portugal, day 140 means July 17. The shaded areas indicate 95% confidence intervals around the predicted model. Actual observations are juxtaposed to the models (•). The R^2^ goodness-of-fit statistic means quadratic Spearman’s *Rho.*

Even though Korean data validates the model, caution is mandatory when interpreting the Portuguese model, as data are incomplete and model parameters are sure to change in time, either accelerating or slowing recovery, depending on the success of mitigation measures and on how the loosing of confinement policies, projected to begin in May, will proceed. It is clear, though, that recovery will be long. With 50% cases still active by July, the risks of new peaks are high, furthermore considering the high percentage of untraced asymptomatic carriers of COVID-19 (Yu et al., 2020). To this we must add the fact that persistence of the virus increases the odds of mutation.

## Data Availability

All data is provided as Supplementary Material.

## Acknowledgements

This work was supported by the Applied Molecular Biosciences Unit (UCIBIO) which is financed by national funds from ‘Fundação para a Ciência e a Tecnologia’, FCT (UIDB/04378/2020). AM is supported by CEEC/IND/00175/2017 contract, BioISI (UIDB/04046/2020 and UIDP/04046/2020) and Green-IT (UID/Multi/04551/2013) grants from FCT.

